# Artificial Intelligence in Healthcare: 2025 Year in Review

**DOI:** 10.64898/2026.02.23.26346888

**Authors:** Ruthvik Edara, Avneesh Khare, Aarit Atreja, Raghav Awasthi, Briana Highum, Natalia Hakimzadeh, Sai Prasad Ramachandran, Shreya Mishra, Dwarikanath Mahapatra, Shalini Shree, Anirban Bhattacharyya, Nishant Singh, Sandeep Reddy, Jacek B. Cywinski, Ashish K. Khanna, Kamal Maheshwari, Francis A. Papay, Piyush Mathur

**Affiliations:** BrainXAI ReSearch, BrainX LLC

## Abstract

**Background:** Breakthroughs in model architecture and the availability of data are driving transformational artificial intelligence in healthcare research at an exponential rate. The shift in use of model types can be attributed to multimodal properties of the Foundation Models, better reflecting the inherently diverse nature of clinical data and the advancing model implementation capabilities. Overall, the field is maturing from exploratory development towards application in real-world evaluation and implementation, spanning both Generative and predictive AI.

**Methods:** Database search in PubMed was performed using the terms “machine learning” or “artificial intelligence” and “2025”, with the search restricted to English-language human-subject research. A BERT-based deep learning classifier, pre-trained and validated on manually labeled data, assessed publication maturity. Five reviewers then manually annotated publications for healthcare specialty, data type, and model type. Systematic reviews, duplicates, pre-prints, robotic surgery studies, and non-human research publications were excluded. Publications employing foundation models were further analyzed for their areas of application and use cases.

**Results:** The PubMed search yielded 49,394 publications, a near-doubling from 28,180 in 2024, of which 3,366 were classified as mature. 2,966 were included in the final analysis after exclusions, compared to 1946 in 2024. Imaging remained the dominant specialty (976 publications), followed by Administrative (277) and General (251). Traditional text-based LLMs (1,019) led model usage, but Multimodal Foundation Models surged from 25 publications in 2024 to 144 in 2025, and Deep Learning models also increased substantially (910). For the first time, publications related to classical Machine Learning model use declined (173) in our annual review. Image remained the predominant data type (53.9%), followed by text (38.2%), with a notable increase in audio (1.2%) coinciding with the adoption of multimodal models. Across foundation model publications, Imaging (110), Head and Neck (92), Surgery (64), Oncology (55), and Ophthalmology (49) were leading specialties, while Administrative and Education categories remained high-volume contributors driven predominantly by LLM-based research.

**Conclusion:** 2025 signals a meaningful maturation of the healthcare AI research field, with publication volumes nearly doubling, classical ML yielding to higher-capacity foundation models, and the field rapidly moving beyond traditional text-based LLM capabilities toward multimodal models. While Imaging continues to lead in research output, the growth of multimodal models across clinical specialties suggests the field is approaching an inflection point where AI systems can more closely mirror the complexity of real-world clinical practice.

## INTRODUCTION

Artificial Intelligence (AI) research in healthcare continues to expand rapidly, and publication volumes reflect sustained growth over time [1]. Building on the structural changes triggered by the introduction of large language models, the model landscape has evolved, with foundation models becoming an increasingly important area of exploration in healthcare AI research [2]. Early enthusiasm focused largely on exploratory use of text-based models across specialties; however, there is a gradual transition toward more deliberate, use-case–driven real-world applications of both generative and predictive AI [3]. Interest in Multimodal Foundation Models seems to be increasing, reflecting the inherently multimodal nature of healthcare data and the need to integrate diverse data modalities, including text, images, and audio, in delivery of healthcare [4].

This review provides a comprehensive assessment of publications related to AI applications in healthcare for the year 2025, along with a comparison of these trends with those observed in the preceding years. To manage large volumes of research, we continue to employ maturity-based evaluations of the peer reviewed publications using a deep learning model, which performs with a high degree of accuracy[5]. Our analysis, as in previous years, also provides a manual analysis of the data and model types to deliver a comprehensive overview of the selected publications. Given the increasing application of generative AI, this review also offers a quantitative analysis and a scoping review of the publications focusing on the rapidly developing field of generative AI within the healthcare sector. Beyond the analysis of the trends related to AI in healthcare research, along with the dataset, this review predicts innovation poised for future clinical implementation and examines the evaluation of commercially available AI.

## METHODOLOGY

Our search for publications was performed using the National Library of Medicine’s PubMed (**Figure 1**) database using the terms, “machine learning” or “artificial intelligence” and “2025”, restricted to English language and human subject research as of December 31, 2025, on January 1, 2026. Pre-print publications, animal research, research related to robotics without the use of AI, publications without abstracts, publications not in English were excluded. Our search methodology has remained consistent since 2019, which allows for comparative analysis of publications for each medical specialty, year over year [2, 3].

**Figure 1:**
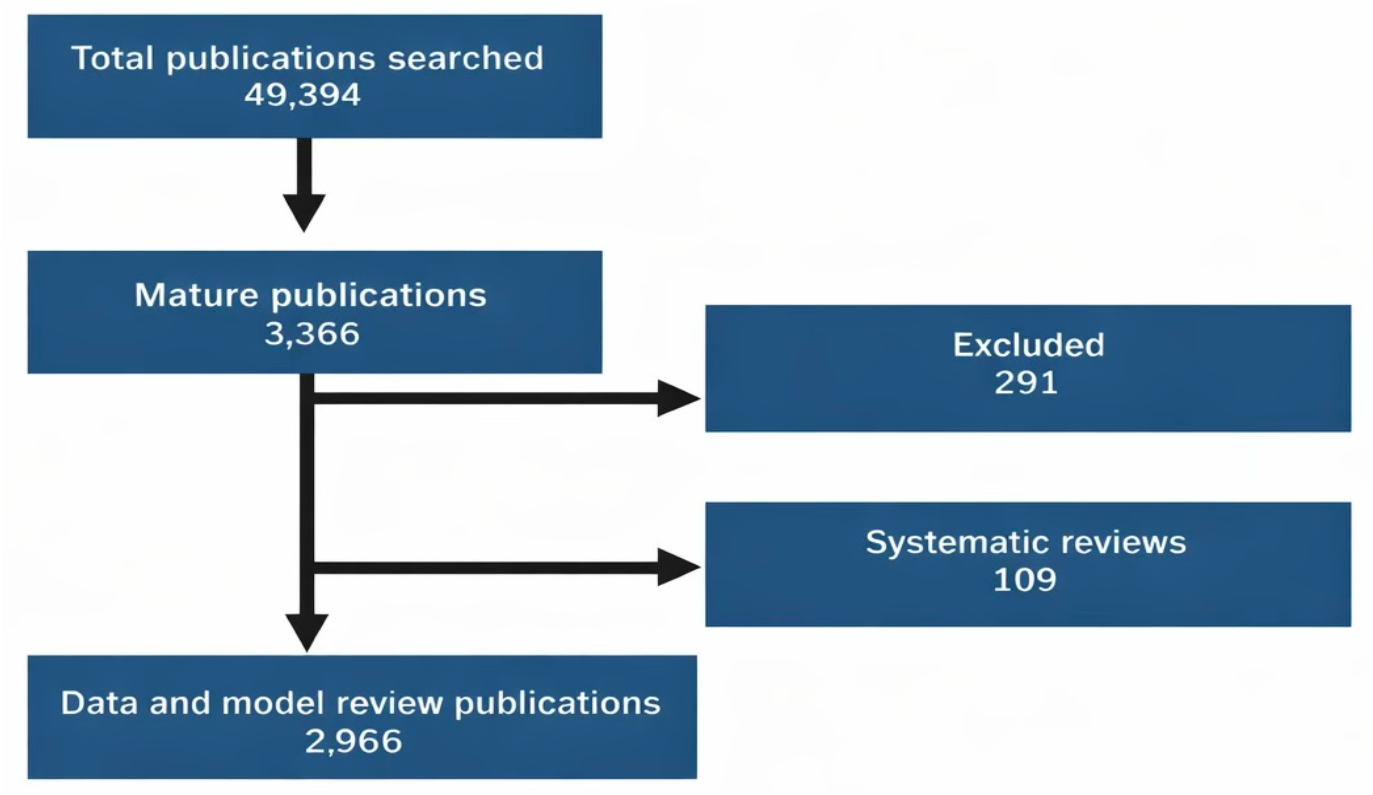
Screening and data extraction for AI in healthcare publications

We used a Bidirectional Encoder Representation from Transformer (BERT)-based maturity classification model that was pre-trained and validated on manually labeled data for ‘Mature’ and ‘Not Mature’ publications [5], to assess the level of maturity of each publication. The level of maturity was determined by the ability of the publication to answer the question: “Does the output of the proposed model have a direct, actionable impact on patient care by providing information to healthcare providers or automated systems?” Systematic reviews were excluded from the count of “mature” publications, as they do not independently address the above question, and the data type and model type are variable in the systematic reviews.

Identified mature publications were manually reviewed, of which 291 publications were excluded from the 3366 mature ones. Excluded publications mostly included review articles, publications related to robotic surgeries or non-human studies. 109 systematic reviews were excluded from the count of “mature” publications.

Further mature publications were classified into 25 categories with 22 healthcare specialities and one General category. Education and Administrative were used as two additional categories where publications specifically focused on these two tasks were categorized. The General category contains many of the publications related to general AI topics that were not specialty-specific, for instance drug development-related publications.

Similar to prior years, five reviewers manually reviewed and annotated the remaining mature publications for the following: healthcare speciality category, data type & model type [2]. Data type was classified manually into four categories of data: image, text, tabular, and voice. Waveform data was categorized within the image data type. Model type was also manually curated from abstracts into eight different categories: Traditional Deep Learning (DL), Classical Machine Learning (ML), AI General, Statistical, Natural Language Processing (NLP), Large Language Model (LLM), Large Vision Model (FM-LVM), and Multimodal Model (FM-MM). Those publications where the model type was not described in the title or the abstract were placed in the “AI General” class. Many of these publications were related to validation studies related to proprietary AI models. Similarly, studies using statistical analysis to analyze surveys or the performance of AI models were included in the statistical class. Due to the significant increase in publications related to foundation models since 2024, we report and provide an analysis of only mature publications using foundation models and its key classes of LLM, FM-LVM, and FM-MM [2].

## RESULTS

Our consistent search methodology continues to show a substantial increase in the number of publications (49,394) related to AI in healthcare in 2025 (**Figure 2A**). Near doubling of the searched publication number and the quantitatively assessed mature publications (3366) using the BERT models once again demonstrates a high number of experiments being performed in this field by the researchers (**Figure 2B, 3**).

**Figure 2:**
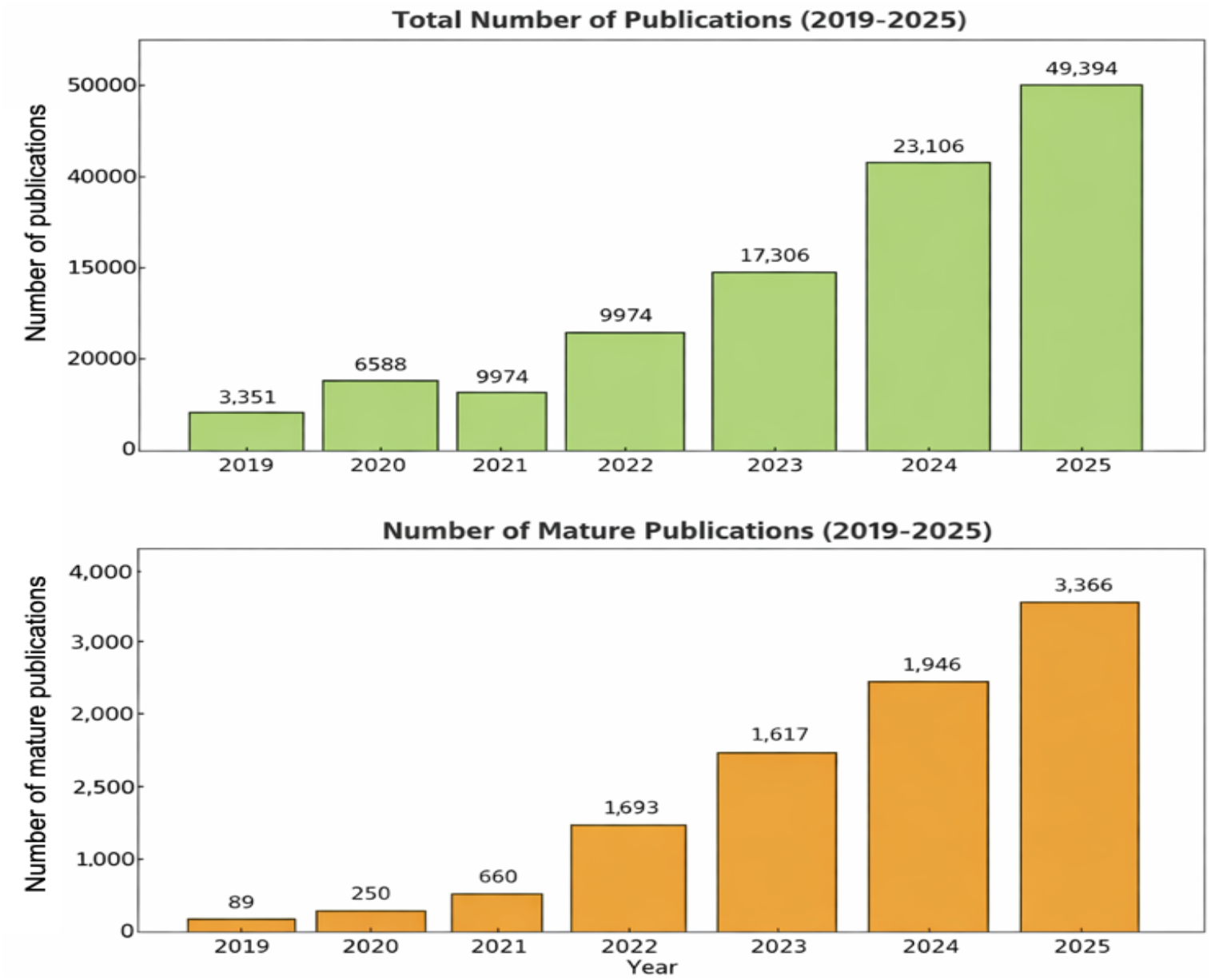
A) Trends in the total number of publications from 2019-2025. B) Trends in the number of mature publications from 2019-2025.

**Figure 3:**
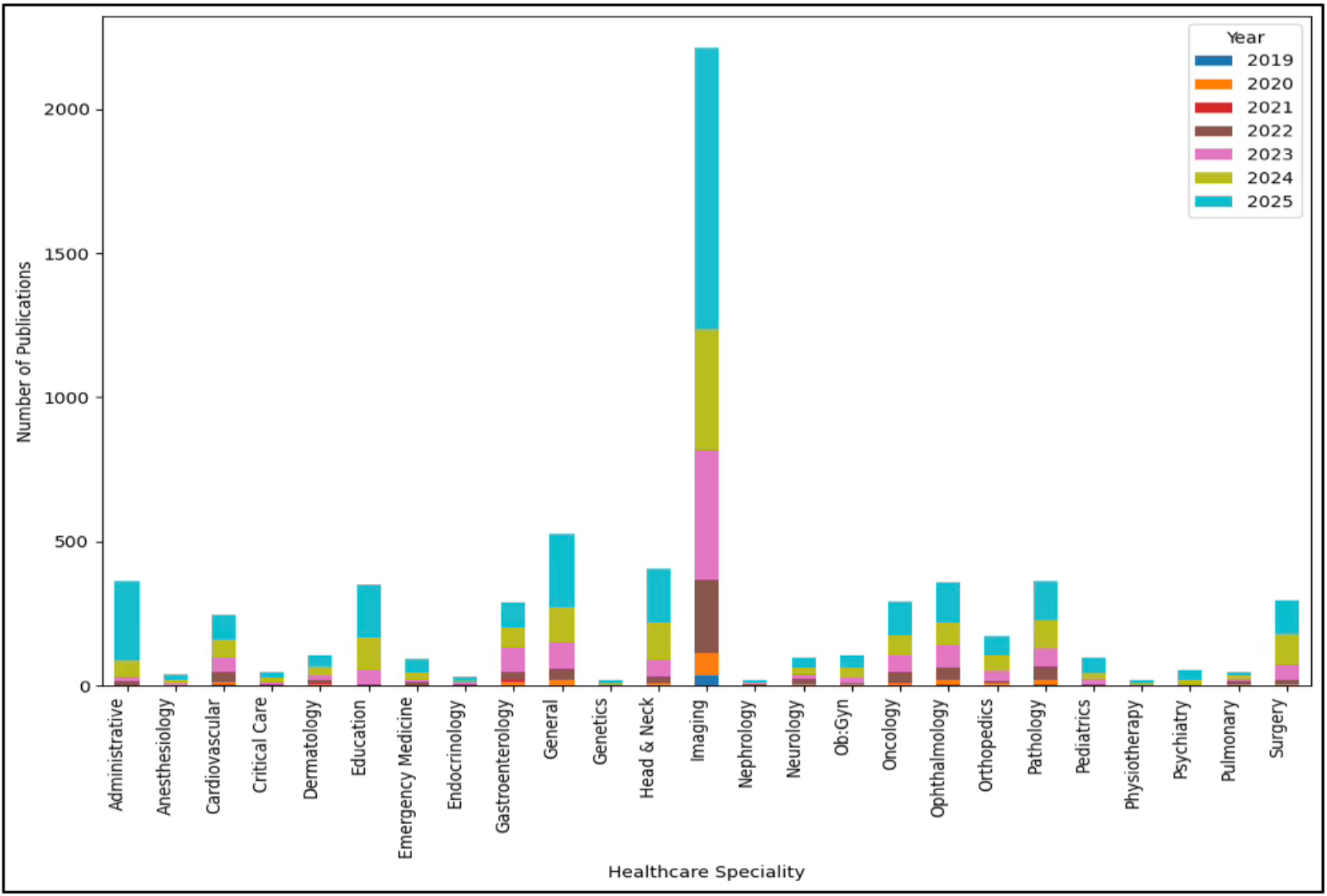
Seven-year distribution in the number of mature publications across different healthcare specialties

Imaging or Radiology (976), as a healthcare speciality, continues to dominate the number of publications this year again along with the Administrative (277), General (251), Head and Neck (185), and Education (181) categories amongst the top five substantial contributors (**Table 1**). Administrative, General and Education as categories rank high taking contributions from all the different specialities and cross speciality application of AI models in healthcare. Ophthalmology, Pathology, Surgery, Oncology, Gastroenterology are additional healthcare specialities which continue to rank high in the number of mature publications generated this year (**Table 1**).

**Table 1:**
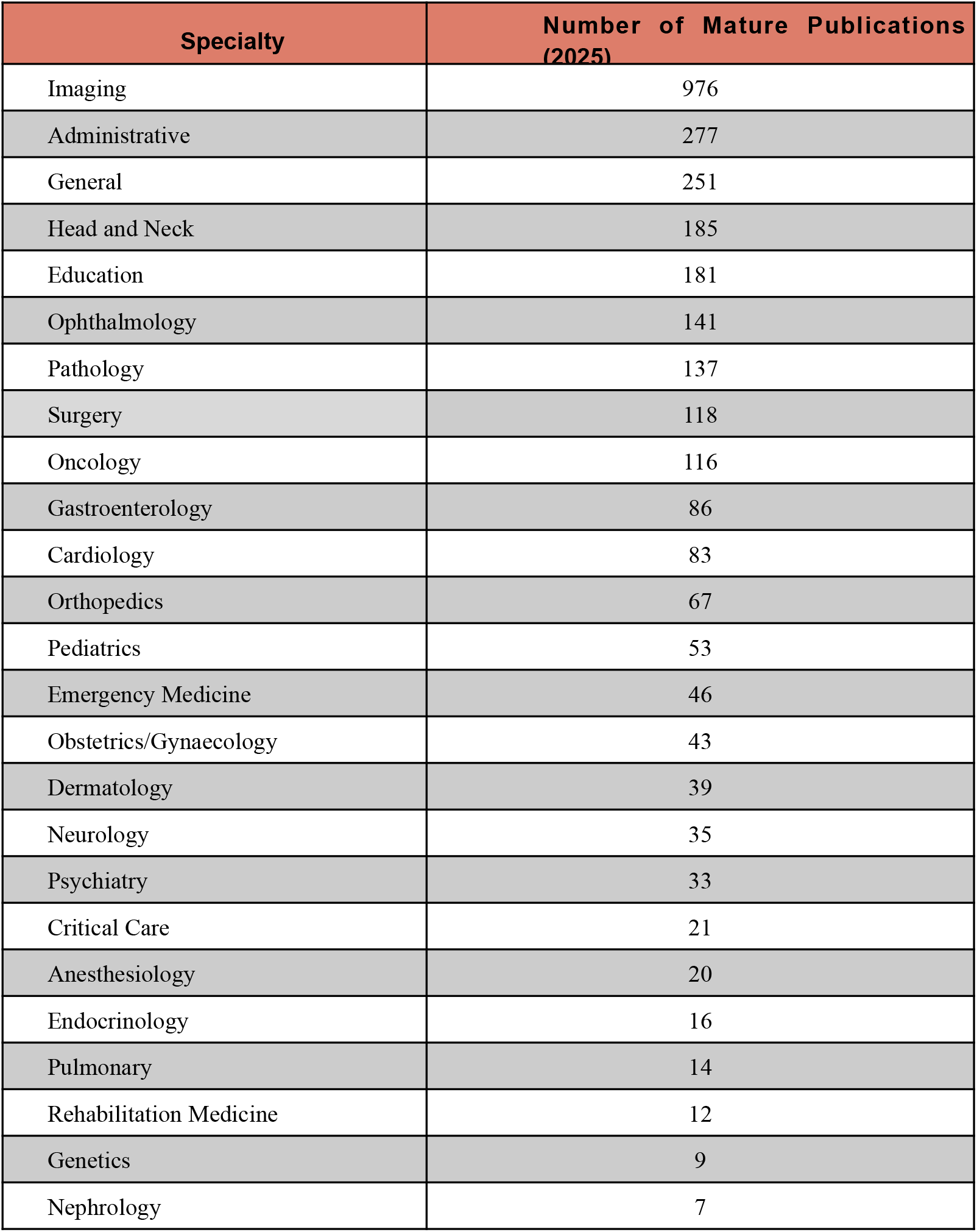
Healthcare specialty mature publication trends for the year 2025.

The breakthrough trend of foundation models as a leading category of models used for AI in healthcare, as seen for the first time in 2024, was preserved once again this year amongst the mature publications (**Figure 4**). Traditional LLMs (1019) continue to lead the model use, but with the evolution of the foundation models towards multimodality, we see a substantial increase in the use of Multimodal Model (FM-MM = 144) and Large Vision Models (LVM = 14) AI in healthcare research (**Figure 4**). Although the use of Deep Learning models (910) and traditional NLP (28) also increased significantly, for the first time we observe a slight decrease in the use of classical ML models (173) compared to last year (**Figure 4**).

**Figure 4:**
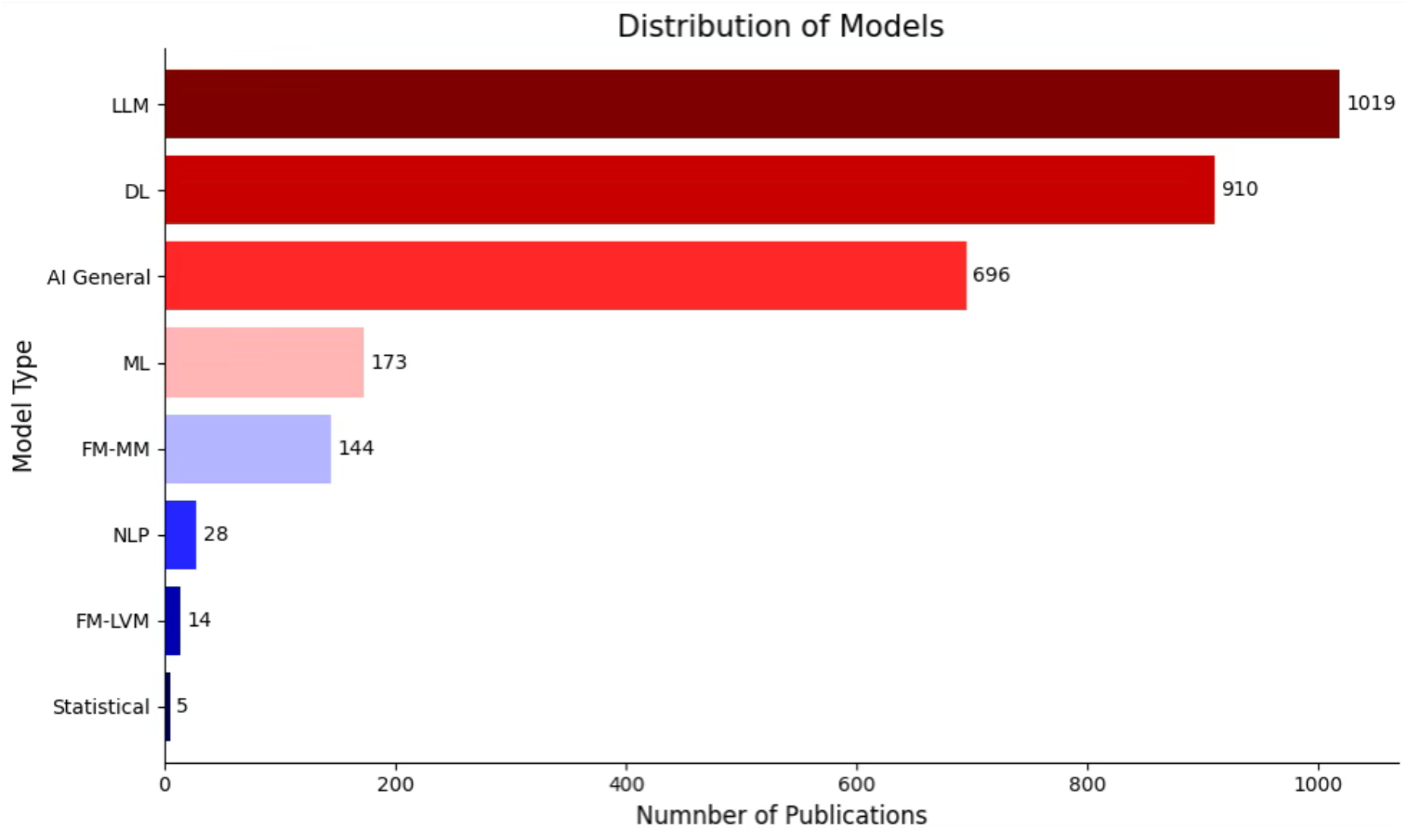
Distribution of model types utilized in the mature articles in the year 2025 (DL = Deep Learning, ML = Machine Learning, LLM = Large Language Model, NLP = Natural Language Processing, FM-LVM = Large Vision Model, FM-MM = Multimodal model)

Compared to 2024, the type of data used amongst the mature AI in healthcare research publications has a similar trend with image (53.9%) and text (38.2%) dominating the data categories (**Figure 5**). Use of multimodal models has probably led to an increase in audio data (1.2%) in 2025 (**Figure 5**). 94 mature publications (3.1%) combined data types, with image and text used together in 54 of these publications, probably due to increasing use of multimodal foundation models. If a publication used more than one data type, credit was given to each of the data categories in our analysis.

**Figure 5:**
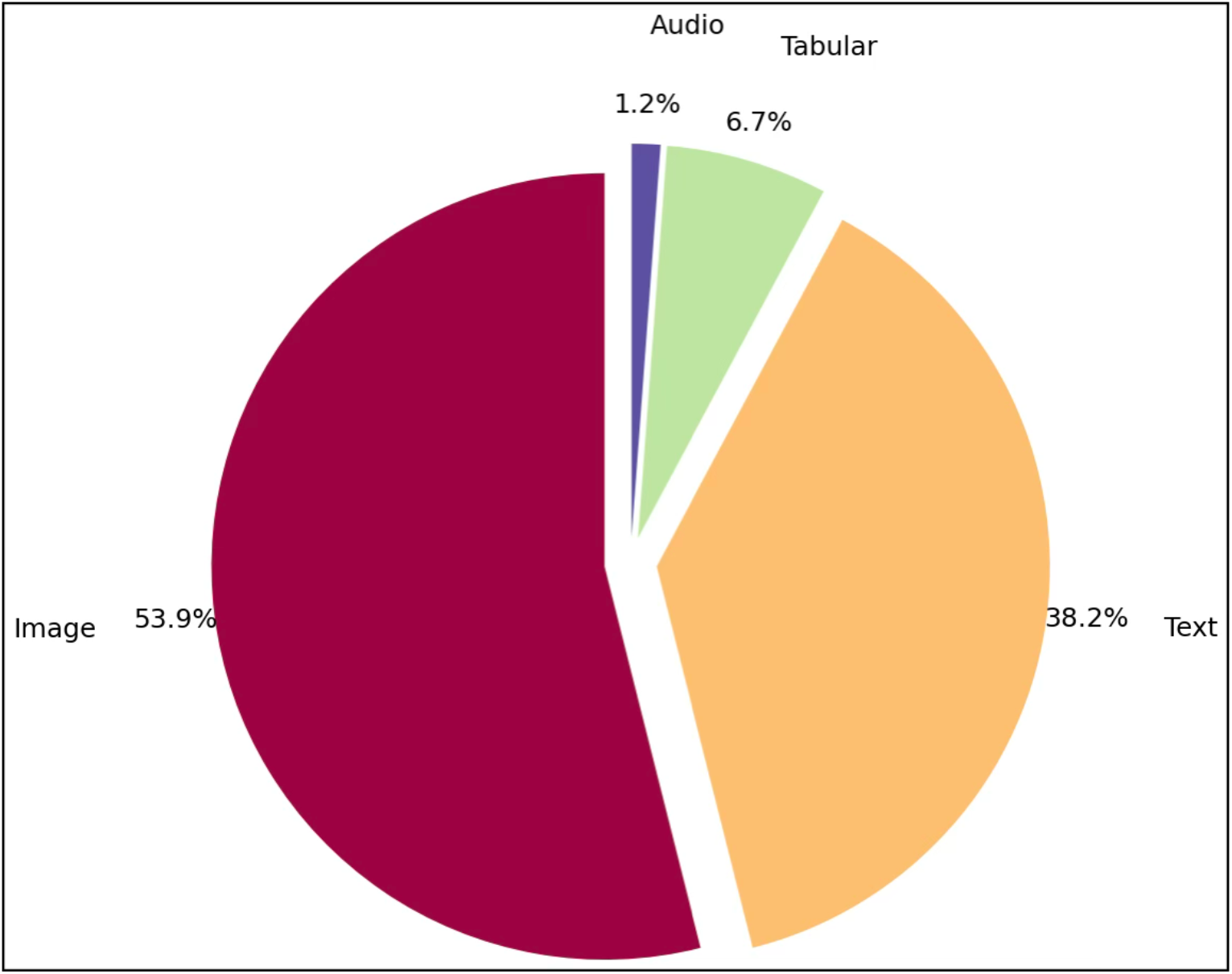
Distribution of data types utilized in the mature publications for the year 2025

Amongst the foundation models, mature publications related to Administrative tasks, Education and General use cases in healthcare continue to dominate (**Figure 6**). Similar to last year, those healthcare specialities that have led the AI in healthcare research and publications continue to lead. Imaging (110), Head and Neck (92), Surgery (64), Oncology (55), and Ophthalmology (49) are notable amongst them (**Figure 6**). This year, in this analysis, we provide a combined data of use of all the foundation models, instead of LLMs only, since the use of multimodal models has substantially increased compared to prior years (**Figure 6**).

**Figure 6:**
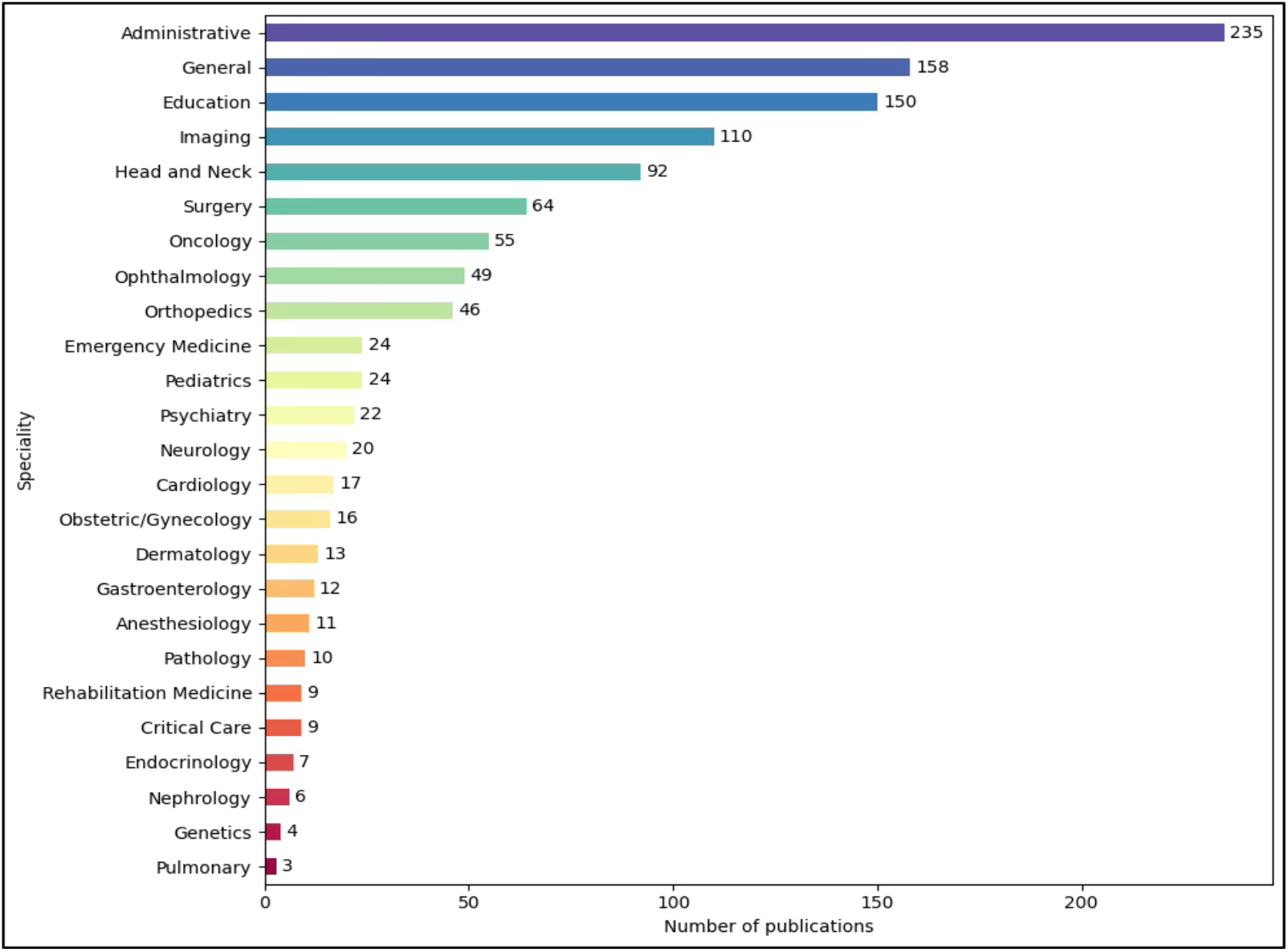
Trends amongst publications based on Foundation Models across healthcare specialties in 2025

## DISCUSSION

AI in healthcare publications continues to grow at a sustained pace in both the quantity and quality of publications. Similar to the shift seen in 2024, we observed a significant evolution in 2025 regarding the complexity of models used. While 2024 was defined by the rapid adoption of text-based Large Language Models (LLMs), 2025 demonstrates a shift toward Multimodal Foundation Models (FM-MM) and a sustained high volume of Deep Learning (DL) research. The substantial increase in mature publications suggests that the field is moving from preliminary exploration to more robust validation [6-8], with a notable increase in publications combining multiple data types to address clinical questions [9, 10].

Domination of Imaging as a healthcare specialty continues with the highest number of publications and continued growth related to AI in healthcare research. Probably based on the significant research community with interest in this field, availability of data, and experience using AI models, Imaging has expanded the use of models from predominantly traditional DL to newer foundation models, including the multimodal models (Figure 4)[11]. These use cases include image interpretation, generation of radiology reports, and summarization of radiology reports, amongst others [12, 13]. Task-based use across healthcare specialities, such as for Education and Administrative tasks, grouped under “specialities”, continues to grow rapidly, predominantly driven by foundation models, using text data. These task-oriented use cases, such as generating patient education material, developing questions for exams, improving and supporting documentation needs, are varied but many times applicable to many of the healthcare specialties [14-16]. The next leading group of healthcare specialities including Ophthalmology, Head and Neck, Pathology, Gastroenterology, Surgery, Cardiology, and Oncology, have also continued to expand their research horizons, expanding their research based on image-based traditional DL models to using foundation models for additional applications in 2025. Image-based dentistry research, grouped within Head and Neck, deserves special mention both for research oriented towards use of image based DL models for use cases such as cephalometry, implant assessment, and the use of foundation models for text based summaries [17-19]. Use of DL for image guidance and therapy determination for radiation therapy is also notable within the speciality of Oncology [20, 21]. While other specialties continue to increase their research efforts, there is once again a lot to be learnt from the leading healthcare specialties in this field, and probably a need for development of resources to foster such research.

We evaluated the distribution of AI models used across all mature publications. For the first time, we observed a slight decrease in the number of publications utilizing classical ML models (173), while the traditional DL models (910) and LLMs (1019) saw substantial increases. This trend suggests a preference for higher-capacity models over traditional models for complex healthcare tasks. Additionally, the use of Multimodal Foundation Models (FM-MM) increased significantly from 25 publications in 2024 to 144 in 2025. This indicates a shift toward models capable of processing diverse data inputs simultaneously. The “AI General” category remains large, often reflecting proprietary models[22-24] where the specific architecture is not disclosed in the title or abstract, further emphasizing the role of commercial tools in current research[25-27].

Since 2023, we have observed a trend towards increased use of text and audio data and this year. Use of FMs has clearly spurred excitement, creating a large community of researchers exploring the use of these models over text, images and audio data [28]. Additionally, more resourced specialities such as Imaging, are also exploring the use of multiple modalities of data in various combinations for their use cases.

Research related to foundation models continues to grow exponentially. LLMs remain the most frequently used foundation model, particularly in areas such as Education and Administrative [29-31]. We observed an increase in the application of Large Vision Models (LVM) and Multimodal Models (FM-MM) in clinical specialties such as Imaging [31-33], Surgery [34], and Pathology[35]. This differs from 2024, where foundation model research was largely text-based. The data suggests that clinical specialties are beginning to adopt foundation models that can interpret diagnostic imagery [36, 37], moving beyond the text-generation focus of early LLM research.

Our methodology for this research has remained consistent since 2019 including publication search limited to the Pubmed database. This consistency has provided us with the ability to trend the scope of research in this expansive area of interests to many healthcare researchers. We believe the number and quality of publications based on a high quality single database search of peer-reviewed publications is adequate to provide information related to the key trends and deeper insights. As a limitation, this methodology might have gaps in missing publications possibly retrievable with the use of additional search databases. Similar to the prior years, our BERT-based approach, despite its limitations, remains content-aware, reproducible even with changing factors such as article citation count and journal impact factor, and fine-tunable with new data. Manual annotations and evaluations can introduce subjectivity, but in our trials, it has remained more accurate than using LLMs or other models.

Despite the development and implementation of AI scribes in various healthcare workflows, we observed a limited number of related publications [38, 39]. With their growing adoption, we might see an increase in the number of prospective evaluation, validation and cost-effectiveness trials. Similarly, with the introduction of Agentic AI approaches for implementation of Generative AI, there were limited publications researching their application in healthcare in 2025, which are likely to grow in the future [40, 41].

## CONCLUSION

AI in healthcare research and publications continue to grow exponentially across all healthcare specialities and tasks. Use of foundation models, especially multimodal foundation models, and text as a type of data has scaled significantly over the last two years. Emerging trends show gains in evaluation and validation of proprietary AI models now deployed in the healthcare workplace. Availability of data, changing techniques and emerging implementation frameworks for deployment of foundation models such as Agentic AI are likely to continue to enhance research and publications related to AI in healthcare in the near future.

## DATA AVAILABILITY

Data is available upon request via email to the corresponding author or by contacting us through our website, BrainXAI ReSearch (https://www.brainxai.com/research). Additional related publications and datasets related to AI in Healthcare can be accessed from BrainX Community website(https://www.brainxai.org).

